# Implications of COVID-19 vaccination and public health countermeasures on SARS-CoV-2 variants of concern in Canada: evidence from a spatial hierarchical cluster analysis

**DOI:** 10.1101/2021.06.28.21259629

**Authors:** Daniel A Adeyinka, Cheryl A. Camillo, Wendie Marks, Nazeem Muhajarine

## Abstract

**Background:** The influence of coronavirus disease-2019 (COVID-19) containment measures on variants of concern (VOC) has been understudied in Canada. Our objective was to identify provinces with disproportionate prevalence of VOC relative to COVID-19 mitigation efforts in provinces and territories in Canada.

**Methods:** We analyzed publicly available provincial- and territorial-level data on the prevalence of VOCs in relation to mitigating factors (summarized in three measures: 1. strength of public health countermeasures: stringency index, 2. how much people moved about outside their homes: mobility index, and 3. vaccine intervention: proportion of Canadian population fully vaccinated). Using spatial agglomerative hierarchical cluster analysis (unsupervised machine learning), the provinces and territories were grouped into clusters by stringency index, mobility index and full vaccine coverage. Kruskal-Wallis test was used to determine the differences in the prevalence of VOC (Alpha, or B.1.1.7, Beta, or B.1.351, Gamma, or P.1, and Delta, or B.1.617.2 variants) between the clusters.

**Results:** Three clusters of vaccine uptake and countermeasures were identified. Cluster 1 consisted of the three Canadian territories, and characterized by higher degree of vaccine deployment and lesser degree of countermeasures. Cluster 2 (located in Central Canada and Atlantic region) was typified by lesser implementation of vaccine deployment and moderate countermeasures. The third cluster was formed by provinces in the Pacific region, Central Canada, and Prairie region, with moderate vaccine deployment but stronger countermeasures. The overall and variant-specific prevalence were significantly different across the clusters.

**Interpretation:** This study found that implementation of COVID-19 public health measures varied across the provinces and territories. Considering the high prevalence of VOCs in Canada, completing the second dose of COVID-19 vaccine in a timely manner is crucial.

## Introduction

The devastating impacts of coronavirus disease-2019 (COVID-19) cannot be overemphasized. With an estimated 172 million cases and 4 million deaths (as of June 2, 2021) worldwide,^1^ the pandemic is unarguably one of the worst in human history. In Canada alone, about 1.4 million people have been infected with severe acute respiratory syndrome coronavirus 2 (SARS-CoV-2)—representing 4% of Canadian residents.^2^ The case fatality rate for COVID-19 in Canada is 2% (i.e., 26,000 deaths).^2^

At this stage of the pandemic, however, there are three forces in ‘tension’ that are vying for prominence: vaccine uptake, variants of SARS-CoV-2 and re-opening of society. In the second year of the pandemic, the emergence and spread of variants of concern (VOCs), concomitantly with rollout of vaccine campaigns, have changed the complexion of the pandemic. Variants, as expected, have added complexity to the nature of COVID-19. Following a slow start to vaccine rollout, uptake has been accelerating in Canada, and many provinces and territories are eager to relax public health countermeasures. However, public health experts remain uneasy about variants-led surges and outbreaks, necessitating reapplication of countermeasures.

To date, four phylogenetic VOCs declared by the World Health Organization (WHO) are being tracked in Canada. The Alpha (B.1.1.7), Beta (B.1.351), Gamma (P.1) and Delta (B.1.617.2) variants, first detected in the United Kingdom, South Africa, Brazil and India, respectively, are been transmitted in Canada and in rest of the world.^3^ Canada confirmed its first cases of Alpha variant in a couple from Toronto who had contact with a traveller from the United Kingdom on December 26, 2020.^3^ The Beta variant was first reported in Alberta on January 8, 2021.^3^ The Gamma variant was first confirmed in Ontario in an international traveller from Brazil on February 8, 2021.^3^ On April 4, 2021, the Delta variant was first reported in British Columbia.^3^

While much still remains to be learned about the epidemiology, diagnosis, management and sequalae of these four VOCs, using the data currently available, we need to understand the spread of VOCs in Canada and how this might be held in check by vaccine uptake rates and public health interventions and countermeasures. Most studies on VOC are either laboratory or clinically based at this time, addressing the efficacy of vaccines on the mutant strains. There is a paucity of population-level studies that assessed the influence of government containment policies and other non-pharmaceutical efforts on VOCs to complement lab-based studies. The knowledge base on VOCs is rapidly emerging and notable national and international initiatives have commenced recently to address this dynamic aspect of COVID-9. In Canada’s decentralized federation, provinces and territories are largely responsible for vaccination and public health responses. To provide insights into the differences in the subnational responses and to support timely government interventions, both at federal, and provincial/territorial levels, this study aims to examine: a) clustering patterns of COVID-19 mitigating efforts, and b) cluster differences in the prevalence of SARS-CoV-2 variants of concern in Canada.

## Methods

### Data sources

We analyzed provincial- and territorial level data on the SARS-CoV-2 VOCs in Canada along with data on mitigating strategies for COVID-19 from publicly available data sources. The outcome variable—prevalence of VOCs by type and total—was estimated as the proportion of cases with VOC per 1 million population, as of June 15, 2021. The cumulative number of VOC cases for Alpha, Beta, Gamma and Delta variants were extracted from a COVID-19 variants of concern tracker in Canada.^4^ Populations at risk were predefined as the first quarter provincial population estimates obtained from Statistics Canada.^5^ The independent variables were layered into vaccine response (i.e., percentage of Canadian population fully vaccinated against COVID-19), policy response (i.e., stringency index—full and partial lockdowns) and behavioural changes (i.e., mobility index).

During the study period, three vaccines (i.e., AstraZeneca, Moderna, and Pfizer-BioNTech) were authorized and in use in vaccination campaigns in Canada. The full vaccination coverage rates were retrieved from the COVID-19 Tracker Canada.^6^ Compared to first dose, full vaccination (two-dose) was selected because the second dose is widely believed to offer greater protection than partial vaccination in slowing down COVID-19 transmission, hospitalizations, disease sequalae and fatalities.^7^

The stringency index is a composite measure generated from 20 indicators such as school and workplace closures, restrictions on public transport, cancellation of public events and gatherings, stay-at-home policies, travel restrictions, public information campaigns, testing policies, contact tracing, and face coverings. These were defined and precalculated by the University of Oxford to track government responses to the COVID-19 pandemic. The policy index was publicly available at the Oxford COVID-19 Government Response Tracker (OxCGRT) website.^8^ The lowest possible score is 0 (mildest) and highest score is 100 (strictest). It should however be noted that stringency index is not a measure of effectiveness of COVID-19 response of government policies, but rather a measure of the degree and comprehensiveness of governmental response, taking account of flexibility of lockdown measures.

To assess individuals’ compliance with government stringency measures (especially reduction in human movement to slow the spread of the virus), we obtained mobility reports from Google LLC.^9^ Through Google Maps app, daily anonymized data on people’s movement in places such as retail and recreation, grocery stores and pharmacies, parks, transit stations, workplaces, and residential areas were collected from smartphones and compared to the baseline five-week period (January 3 to February 6, 2020)—pre-pandemic period. To determine mobility pattern across the provinces/territories, we estimated the average number of visits to each category of places visited between January 1, 2021 and June 15, 2021. The average of the mobility patterns across the entire study period was estimated to account for changes in movement due to weather and holidays, and also any within-province variations of public health countermeasures. Using the first component of principal components analysis (PCA), the dimensions of the six variables that measured changes in movement of people relative to pre-COVID era were reduced with singular value decomposition (SVD) method and z-score transformation (i.e., mean of zero and variance of one). The first component (interpreted as mobility index) explained 60% of the total variance and its eigenvalue was 3.60. The first component had positive loadings on residential areas (0.47) and parks (0.36), but negative loadings on workplace (−0.5), transit stations (−0.49), grocery stores (−0.11) and retail/recreational centres (−0.39). For each variable, a positive loading suggests higher mobility index, and negative loading indicates lower mobility index.

### Statistical analysis

We conducted a spatial agglomerative hierarchical cluster analysis (unsupervised machine learning) to detect clusters of spatial (dis)similarities in COVID-19 mitigating factors in GeoDa™ version 1.18 software.^10^ Furthermore, we determined difference in prevalence of VOCs across the clusters. A symmetrical distance-based weight matrix with an optimal arc distance of 7000km was generated. After many calibrations, single-linkage clustering with Euclidean distance function and geometric centroid weight of 1 was considered appropriate and used.

A spatial cluster analysis was performed using the mitigating factors (i.e., stringency index, mobility index and full vaccine coverage). With the single-linkage, inter-cluster distance was determined by the closest distance between the observations (i.e., closest neighbour clustering). The first step was to transform the independent variables using z-standardization since they were in different scales. In an attempt to select the number of clusters that provided the best fit (distinct clustering), we used stopping rule—Duda-Hart index. We selected groups with the highest Duda-Hart index (0.64) and lowest pseudo T-squared (4.42). The stopping rule corresponds to three clusters reported.

Kruskal-Wallis equality-of-population rank test was used to determine the differences in the prevalence of SARS-CoV-2 variants of concern among the clusters in Stata™ version 17.0 software.^11^ Post-hoc pairwise (posteriori) comparisons of the clusters were performed with Dunn’s test; false detection rates (FDRs) were minimized by using Benjamini-Hochberg adjustment. The statistical significance was set at two-sided p<0.05. To visualize the relationship between the prevalence of VOC, vaccine uptake and countermeasures, bivariate choropleth maps were generated in QGIS™ version 3.12.1 software.^12^ The bivariate maps were based on quantile classification (i.e., tertile), see Figures 2-4. As shown, the 3 by 3 two-dimensional color palette density becomes progressively darker as it moves from lower to higher tertiles. We report the observed value ranges for each tertile classification.

### Ethical approval

This analysis centred on publicly available data with no identifiable information about the people studied. Therefore, research ethics board approval was not required for this study.

## Results

Table 1 shows the distribution of SARS-CoV-2 VOC in Canada. As of June 15, 2021, four VOCs (Alpha, B.1.1.7; Beta, B.1.351; Gamma, P.1 and Delta, B.1.617.2) have been identified, for a prevalence of 6157.47 per 1 million population and 16.69% of all cases. Nova Scotia reporting lowest at 89.85 per 1 million population to Alberta the highest at 10,848.11 per 1 million population. At 91.4% of all VOCs, Alpha variant was the predominant strain in Canada. The Gamma variant accounted for 6.53%, Beta variant, 0.82%, and Delta variant, 0.82% of the mutant strains. While Alberta and Ontario residents had higher prevalence of Alpha variant, lower prevalence of this strain was observed in Yukon and Nova Scotia. The prevalence of Beta variant was highest in Ontario, followed by Quebec, however, Gamma and Delta variants were more common in British Columbia and Alberta.

Compared to the baseline (January 3, 2020-February 6, 2020), mobility related to home/residential areas increased by 12.74% between January 1, 2021 and June 15, 2021 among Canadians. On average, movement of people to parks and outdoor spaces increased by 38.75%; however, in Prince Edward Island, it decreased (average trend= -45%). Overall, mobility related to visits to grocery/pharmacy stores decreased by 4.62% across Canada, but increased in Nova Scotia (by 2.64%), British Columbia (2.45%) and in Saskatchewan (1.91%). Across Canada, movement related to public transport stations decreased by 56.47%, retail and recreational centres by 29.41% and workplaces by 31.34%.

The national stringency index for COVID-19 was 73.61% (lowest in Yukon at 47.22% and highest in Ontario at 90.74%). The Canadian population fully vaccinated against COVID-19 was 13.77% (lowest in Newfoundland and Labrador at 5.67% and highest in Yukon at 61.56%.)

Table 1: Distribution of SARS-CoV-2 variants of concern in Canada, June 15, 2021

### Spatial hierarchical clustering

Cluster analysis with single-linkage method identified three cluster profiles of VOC prevalence and vaccine uptake and public health countermeasures among Canadian provinces/territories (see Table 2 and Figure 1). The clusters were significantly different from one another in their average prevalence of COVID-19 variant cases and variant-specific prevalence, vaccine uptake and public health countermeasures (see Table 2). Table S1 (Supplementary File) shows the frequency distribution of the variables after Benjamini-Hochberg correction for post-hoc pairwise comparisons.

**Figure 1:**
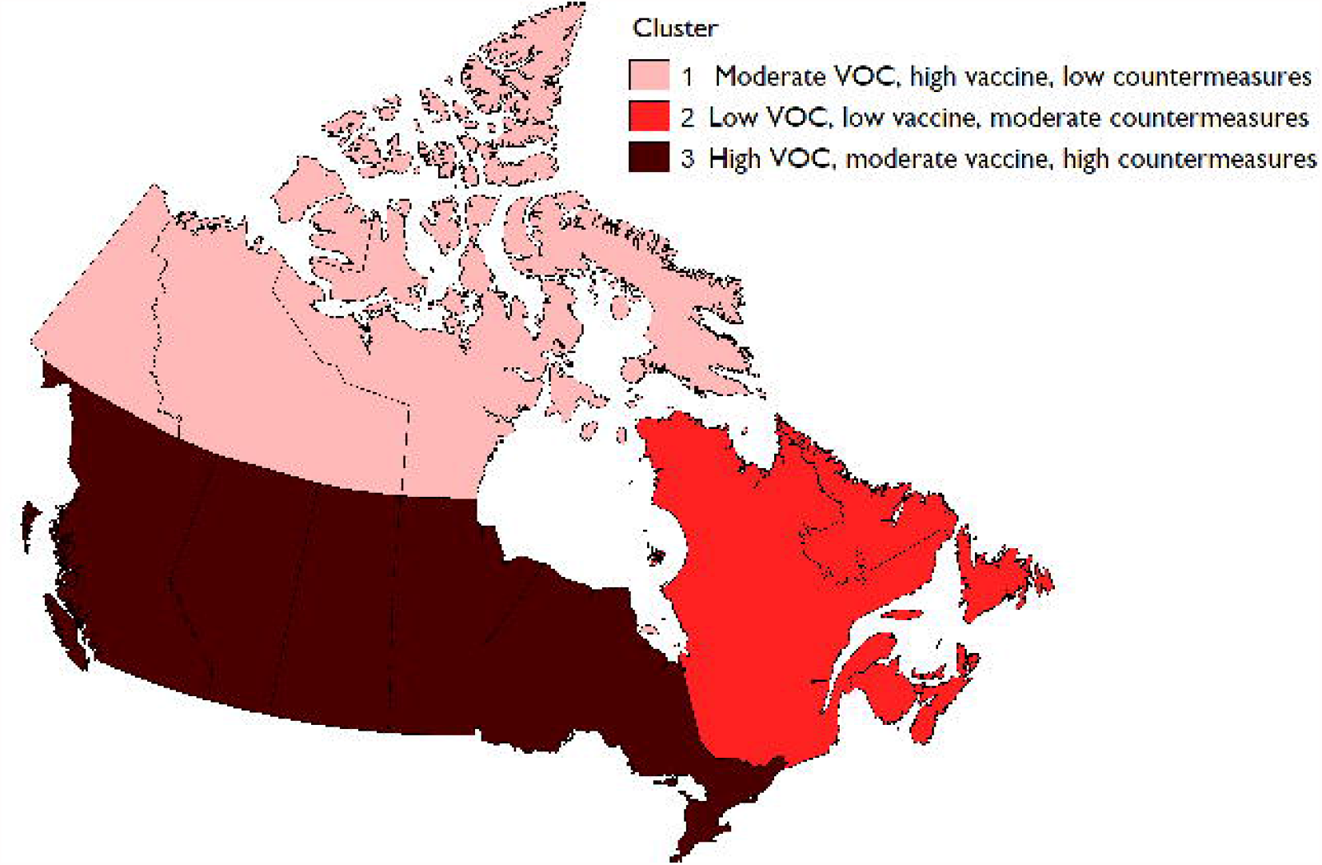
Spatial clusters of prevalence of SARS-CoV-2 variants of concern, vaccine uptake and public health countermeasures in Canada, June 15, 2021.

Yukon, Northwest Territories and Nunavut, the first cluster, is described as having moderate prevalence of VOCs, high vaccine uptake (fully vaccinated), and low countermeasures. The four Atlantic provinces, New Brunswick, Newfoundland and Labrador, Nova Scotia, Prince Edward Island, and Quebec, the second cluster, is described as low prevalence of VOCs, low vaccine uptake (fully vaccinated), and moderate mobility and moderate countermeasures. The four western provinces, British Columbia, Alberta, Saskatchewan, and Manitoba, along with Ontario, the third cluster, is described as having high prevalence of VOCs, moderate vaccine uptake, high mobility, and high countermeasures.

#### Cluster-profile 1 (Yukon, Northwest Territories, and Nunavut, 23% of variance); moderate prevalence of VOCs, high vaccine, low stringency, and low mobility

Compared to other clusters, Northwest Territories, Nunavut and Yukon were characterised by moderate prevalence of aggregated VOC and Alpha variant (533 per 1 million population), and Gamma (22 per 1 million population). It is important to note that the Beta and Delta variants have not been identified in the three territories of cluster 1. In addition, cluster 1 had the highest proportion of Canadians who had received two doses of COVID-19 vaccines (58.53%), lowest mobility index (−2.09), and lowest stringency index (58.49%).

#### Cluster-profile 2 (New Brunswick, Newfoundland and Labrador, Nova Scotia, Prince Edward Island and Quebec, 38.46%); low prevalence of VOCs, low vaccination, moderate stringency, and moderate mobility

Compared to other clusters, cluster 2 was characterised by lowest prevalence of aggregated VOCs (237 per 1 million population), Alpha (230 per 1 million population) and Gamma (1 per 1 million population). However, it had relatively moderate levels of Beta (12 per 1 million population) and Delta (2 per 1 million population). In addition, cluster 2 had lowest full vaccination coverage rates (10.49%), moderate mobility index (0.41), and moderate stringency index (68.61%).

#### Cluster-profile 3 (Alberta, British Columbia, Manitoba, Ontario and Saskatchewan, 38.46%); high prevalence of VOCs, moderate vaccination, high stringency, and high mobility

Compared to other clusters, cluster 3 had the highest prevalence of VOCs (5461 per 1 million population) and variant-specific prevalence—Alpha (5200 per 1 million population), Beta (33 per 1 million population), Gamma (280 per 1 million population) and Delta (73 per 1 million population). Also, cluster 3 had moderate fully vaccinated coverage rates (15.85), highest mobility index (1.25) and highest stringency index (71.57%).

Table 2: Characteristics of clusters from spatial hierarchical clustering analysis of mitigating factors

*\*Inter-cluster differences with Kruskal-Wallis test, IQR: interquartile range, SD: standard deviation, YT: Yukon, NT: Northwest Territories, NU: Nunavut, NB: New Brunswick, NL: Newfoundland and Labrador, NS: Nova Scotia, PE: Prince Edward Island, QC: Quebec, AB: Alberta, BC: British Columbia, MB: Manitoba, ON: Ontario and SK: Saskatchewan*

### SARS-CoV-2 variants of concern and vaccine uptake

Figure 2 shows the distribution of VOCs and the proportion of Canadians who have received the complete schedule of COVID-19 vaccine by province. Provinces shown in the darkest colour (towards top right in the legend in map) have relatively high VOC prevalence and high vaccination rates; those in the lightest colour (bottom left) have low VOC prevalence and low vaccination rates.

**Figure 2:**
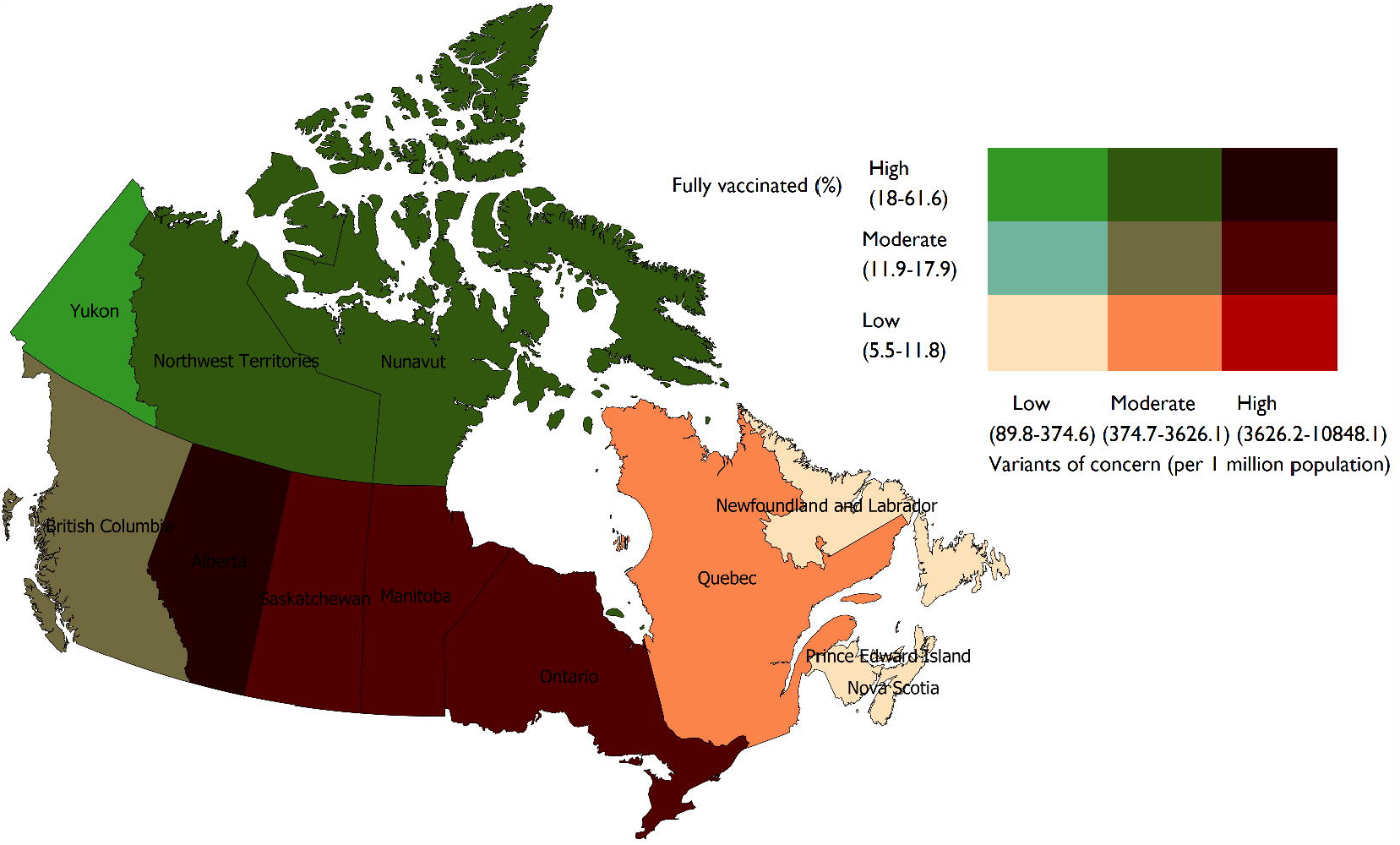
Association between variants of concern and 2-doses of COVID-19 vaccine, Canada, June 15, 2021

Of all provinces and territories, Alberta is classified as marginally higher vaccine rate and high VOC prevalence: 18.89% people fully vaccinated and 10,848 per 1 million population VOC prevalence. Ontario (9,919 per 1 million population), Saskatchewan (5,461 per 1 million population), and Manitoba (4,598 per 1 million population) had high prevalence of VOC and relatively moderate vaccine uptake rates (Ontario: 13.83%, Manitoba: 15.85% and Saskatchewan: 17.98%).

At the opposite end, meaning low VOC prevalence and low vaccine rates, the Atlantic provinces recording low VOC prevalence and low vaccine rates were: Nova Scotia (VOC prevalence 90 per 1 million population and 5.55%), Prince Edward Island (175 per 1 million population and 10.85%), New Brunswick (237 per 1 million population and 10.49%), and Newfoundland and Labrador (374 per 1 million population and 5.67%).

Québec and British Columbia had moderate levels of VOC prevalence and vaccine uptake rates (Quebec, VOC prevalence 900 per 1 million population and vaccine rate 11.9% and British Columbia 3626 per 1 million population and vaccine rate 12.79%). Yukon had low prevalence of VOC (190 per 1 million population) and high vaccine rate (41.56%), followed by Northwest Territories (1706 per 1 million population and 58.53%)) and Nunavut (533 per 1 million population and Nunavut (40.39%).

### SARS-CoV-2 variants of concern and public health countermeasures

Figure 3 shows the relationship between the prevalence of VOC and government stringency measures to curb COVID-19. The provinces of Ontario and Manitoba show higher prevalence of VOCs (9919 and 4598 per 1 million population, respectively) and high score on the stringency index—Ontario (90.74%) and Manitoba (75.93%). Saskatchewan recorded a higher prevalence of VOC (5461 per 1 million population) and lower stringency (57.85%). Alberta had high prevalence of VOC (10,848 per 1 million population) and moderate stringency (68.52%).

**Figure 3:**
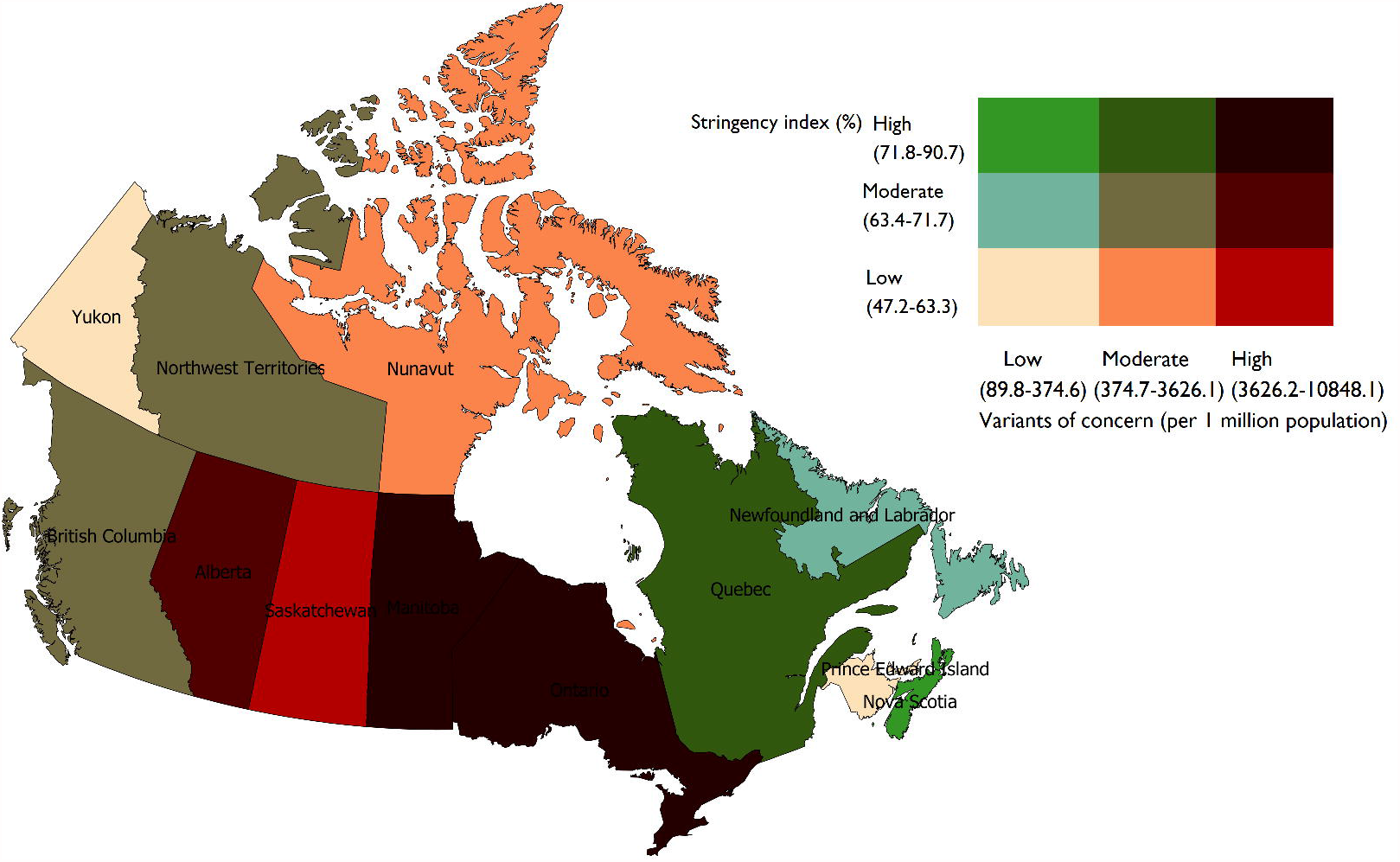
Association between variants of concerns and stringency measures, Canada, June 15, 2021.

The province of Quebec had moderate level of VOC prevalence (900 per 1 million population) with high stringency (73.15%). British Columbia and Northwest Territories have moderate prevalence of VOCs (3626 per 1 million population and 1706 per 1 million population, respectively) and moderate level of stringency—British Columbia (64.81%) and Northwest Territories (64.81%).

At the low-end of VOC prevalence, Yukon, Prince Edward Island and New Brunswick had lower stringency--Yukon 47.22%, Prince Edward Island 58.33% and New Brunswick 58.33%. Nova Scotia reports lower prevalence of VOC (90 per 1 million population) and relatively higher stringency (81.48%), and Newfoundland and Labrador lower prevalence of VOC at 374 per 1 million population and moderate stringency at 71.76%.

### SARS-CoV-2 variants of concern and mobility index

Figure 4 presents the association between prevalence of VOCs and changes in movement of people in relation to the beginning of the pandemic (mobility index). Amongst the provinces reporting higher VOC prevalence, populations in Ontario and Alberta showed higher mobility index— Ontario (3.01) and Alberta (1.31). In this higher prevalence VOC group, Manitoba and Saskatchewan populations had moderate mobility index—1.19 and 0.2, respectively.

**Figure 4:**
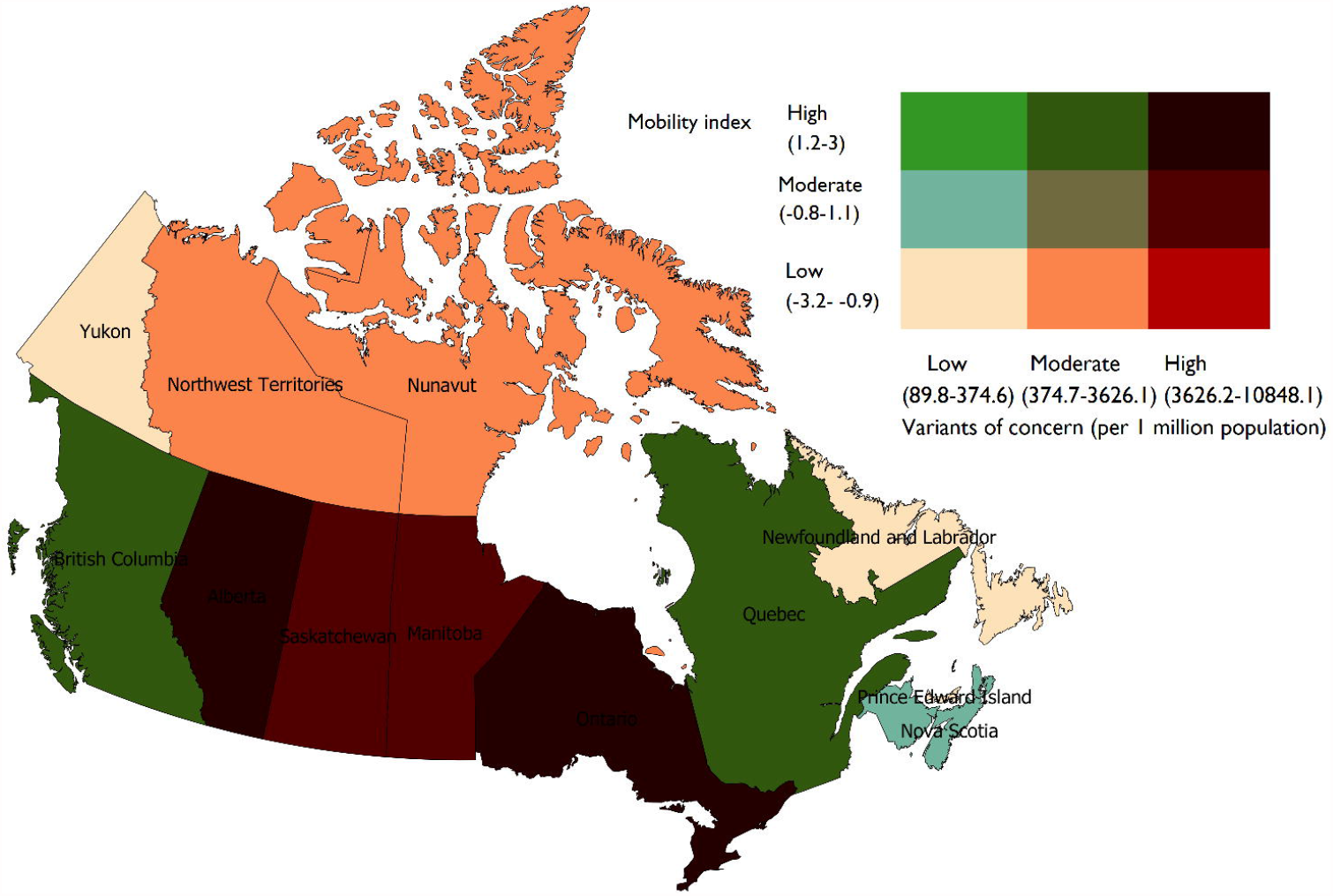
Association between variants of concern and mobility index, June 15, 2021 *Mobility index was derived from the first component of principal components analysis of data on movement of people between Jan 1, 2021 and June 15, 2021, compared to January 3 to February 6, 2020, across different types of places such as retail and recreation, groceries and pharmacies, parks, transit stations, workplaces, and residential*.

Amongst the provinces and territories reporting moderate VOC prevalence, Quebec and British Columbia recorded higher mobility index— Quebec (2.07) and British Columbia (1.25), while Northwest Territories (−1.04) and Nunavut (−3.22) had lower mobility index.

Two of the four Atlantic provinces, Prince Edward Island and Newfoundland and Labrador, and Yukon recorded low VOC prevalence and low mobility index—Prince Edward Island at -2.71, Newfoundland and Labrador -1.04 and Yukon -2.09. Nova Scotia and New Brunswick recorded low prevalence of VOCs and moderate mobility, 0.7 and 0.41, respectively.

### Interpretation

As with most COVID-19 research, this study sought to understand the disease as it is unfolding; specifically, SARS-CoV-2 variants of concern in Canadian provinces and territories at a ‘point-in-time’. By June 15, 2021, the daily new case rate in all Canadian provinces and territories were declining, indicating the resolution phase of the third wave of the epidemic curve. At the same time, however, much concern is expressed that the VOCs, in particular the Delta variant, could trigger widespread outbreaks, especially alongst the unvaccinated or partially vaccinated. Many provinces have set in motions plans to relax public health countermeasures, relying on vaccination rates as the criterion for re-opening.

Across Canadian provinces and territories, we have shown a pattern of VOC spread, vaccine uptake (both doses) and governmental countermeasures that can be profiled in three clusters. The first cluster, comprising all three Canadian territories, is characterized by moderate VOC prevalence, higher degree of vaccine uptake and lesser degree of governmental countermeasures. The second cluster, comprising the Atlantic provinces of Nova Scotia, Prince Edward Island, Newfoundland and Labrador and New Brunswick, along with the province of Quebec is characterized by low VOC prevalence, low vaccine uptake and moderate stringency of governmental countermeasures. The third cluster, comprising Ontario, Manitoba, Saskatchewan, Alberta, and British Columbia, is characterized by high prevalence of VOCs, moderate vaccine uptake and stricter stringency by governmental countermeasures. Strikingly, inter-cluster disparity in the prevalence of VOCs is more evident for cluster three, making the provinces in that group hotspots of VOC.

As governments across Canada continue to enact their plans for easing public health countermeasures, there is concern that Canada needs to have much higher proportion of its population receive both doses of the vaccine. While the per capita case rates are declining across Canada, there have been recent increase in the highly transmissible variant of concern, the Delta variant (which has been shown to be 60% more infectious) in some provinces.^13,14^ These recent regional outbreaks were seen mostly among unvaccinated people.^15^

As government strives in its efforts to get many people vaccinated within the shortest time possible, re-opening plans may need to be further calibrated depending on the local spread of VOCs and vaccine hesitancy and refusals. There have been several alarms raised that the Delta variant could become dominant if people did not complete the full COVID-19 vaccination.^16,17^ In a recent study conducted in Scotland, the first dose was shown not to confer complete immunity against the Delta variant than the Alpha variant, however, full vaccination (two-doses) improves immune effectiveness.^16^ Our present study confirms that in the post-vaccine rollout in Canada (i.e., December 2020), vaccination coverage rates are uneven across the country. While Northwest Territories and Yukon had fully vaccinated more than half of their residents, Newfoundland and Labrador and Nova Scotia had only vaccinated 6% of their residents.

At least three strains of VOC with different transmission risks and responses to COVID-19 vaccines were concomitantly identified in more than half of the provinces. Like in the United States^18^, in the first quarter of 2021, the dominant variant in Canada is Alpha variant and Delta variant is gradually becoming dominant. According to Davies et al.,^18^ the Alpha variant has a high reproductive number (43-90%). However, a more worrying development is the identification of Beta and Delta variant in 77% of the provinces. Due to the key mutations at E484K and K417N receptor-binding sites of the spike proteins for Beta,^19–22^ and L452R receptors for Delta, ^23^ both variants have the tendency to escape recognition by neutralizing antibodies (nAbs), thus evading both natural and vaccine-induced immunity. Also, T478K is not well recognized as responsible for immune evasion yet for Delta variants, and may, like 501 be more relevant for ACE2 binding, however, additional mutation at K417N have been reported for Delta variant. Given the relatively high prevalence of Gamma variant in some provinces/territories (i.e., British Columbia, Alberta, Ontario, Saskatchewan, Manitoba and Yukon), Gamma variant also evades the immune system (but less so than Beta variant) through a significant change in a nAb epitope (E484K and K417T).

This observation has far reaching consequences on health outcomes and prolongation of the epidemic due to new infections. Considering the high prevalence of Beta, Gamma, and Delta variants in some provinces (i.e., Ontario, Quebec, Alberta, British Columbia, Saskatchewan, and Manitoba), stakeholders should continue to emphasize two-dose vaccine uptake and maintenance of public health countermeasures such as mask wearing and social distancing. While the structural and operational barriers to vaccination (e.g., vaccine stock-out, long waiting time and vaccine refusal/hesitancy) need to be tackled, people must be adequately sensitized to complete the second dose to offer full population protection against the VOCs (especially Beta, Gamma, and Delta), thereby reducing future risk of new variants.

The discordance between VOC prevalence and stringency measures warrants cautious interpretation. Our analysis could not show whether the relaxation of lockdown policies which allowed more freedom in the territories in Canada was informed by declining COVID-19 cases and progressive vaccine rollout, or vice-versa. This inverse relationship, which is quite possible, means the timing of implementation of polices in relation to the changing epidemiological contexts of the pandemic is important. At the time of conducting this study, no comprehensive time series data are easily available for Canada. Further investigation with time-series will shed more light on the observed phenomenon. However, we could establish that successful immunization campaigns in the territories in northern Canada have played a critical role in lowering VOC cases. Fast tracking second-dose vaccination is not only key to lessening stringency measures for communities to return to normalcy, but it has been shown in previous studies^24,25^ to curtail transmission of COVID-19 variants.

Our results noted some spatial outliers (i.e., discordant areas) for the relationship between the selected public health measures and VOC in Ontario and in the Canadian Prairie provinces. To optimize public health impacts, these provinces need to revisit some of their approaches and re-prioritize specific interventions. It is also noteworthy to further examine the intra-provincial variations between the public health measures and patterns of VOC for the spatial outliers.

## Limitations

Overall, this study contributes to the current research needs to guide stakeholders on preventive measures to curtail VOCs. Also, the bivariate choropleth map eased readability of spatial patterns, compared to proportional symbol maps. This study has some limitations. The recommendations for administration of different vaccine products (e.g., mRNA based, viral-vector based) have been quite fluid in Canada; we have not taken into consideration the spatial associations between different vaccine products and VOCs. Further research is needed to specifically assess the geographical patterns of vaccine products and VOCs, especially for Beta, Gamma, and Delta variants. As mentioned, this is an ongoing information need, rather than one-time project.

The biggest limitation, however, is the time lag and the uneven testing, detecting, and sequencing efforts to identify VOCs in Canada. Whole Genomic Sequencing (WGS) efforts to identify VOCs by large volumes are currently lagging in Canada. Also, there is selection bias of the samples that are sent for sequencing. The provinces and territories have different criteria for sending samples for sequencing, which could delay detection and bias the proportion of VOCs associated with Beta and Gamma variants. For example, in Ontario, the WGS was triggered from quantitative Polymerase Chain Reaction (qPCR) testing for E48K. The samples with the E48K mutations were prioritized for WGS. However, in May 2021, WGS algorithm changed to randomly selected sampling of 10% positives, with the proportion then increasing to 50% and now to all positives. This calls for new and rapid detection methods for VOCs even as Canada continues to fully vaccinate its populations. Also, there is possibility of systematic bias in Google mobility index being under-estimated among people without smartphones or internet connection (especially in the territories). Due to how the mobility patterns were captured by Google LLC— geographical jurisdiction, it is challenging to delineate international travels and inter-provincial and territorial mobility.

## Conclusion

In conclusion, this study found that SARS-CoV-2 variants of concern in Canadian provinces and territories, to date, show discernible geographical clustering patterns: the territories recording low VOC prevalence, Atlantic provinces and Quebec recording moderate VOC prevalence, and Western provinces and Ontario recording high VOC prevalence. A fuller picture of VOC emerges when its prevalence is correlated with the proportion of populations having received two-doses of vaccines, governmental countermeasures and mobility. Surveillance of VOCs should continue across Canada, while accelerating the rollout of second dose of vaccine and achieving a balance in relation to lifting and relaxation of public health countermeasures.

## Data sharing

Data on cumulative number of VOC cases can be downloaded from CTV News COVID-19 variants of concern tracker,^4^ and vaccine coverage rates are available at the COVID-19 Vaccination Tracker.^6^ Provincial- and territorial-level population estimates are available from Statistics Canada.^5^ Stringency index can be obtained at Oxford COVID-19 Government Response Tracker (OxCGRT) website.^8^ The mobility reports can be downloaded from Google LLC.^9^ Researchers who require access to the study data can contact the corresponding author for further information.

## Data Availability

Data sharing: Data on cumulative number of VOC cases can be downloaded from CTV News COVID-19 variants of concern tracker,4 and vaccine coverage rates are available at the COVID-19 Vaccination Tracker. Provincial- and territorial-level population estimates are available from Statistics Canada. Stringency index can be obtained at Oxford COVID-19 Government Response Tracker (OxCGRT) website. The mobility reports can be downloaded from Google
LLC. Researchers who require access to the study data can contact the corresponding author
for further information.

## Acknowledgements

The authors would like to acknowledge the valuable feedback on previous drafts provided by colleagues in CIHR Coronavirus Variants Rapid Response Network, CoVaRR-Net, specifically Dr Anne-Claude Gingras (Lunenfeld-Tanenbaum Research Institute, Mount Sinai Hospital, Sinai Health and Department of Molecular Genetics, University of Toronto) and Dr Marc-Andre Langlois (Director of the University of Ottawa Biocontainment Facility and Department of Biochemistry, Microbiology and Immunology, University of Ottawa).

